# Combined Exercise Training vs Health Education for Older Adults with Hypertension: The HAEL Randomized Clinical Trial

**DOI:** 10.1101/2025.10.14.25337942

**Authors:** Lucas Porto Santos, Cíntia Ehlers Botton, Eurico Nestor Wilhelm, Nórton Luís Oliveira, Gustavo Zaccaria Schaun, Cristine Lima Alberton, Gustavo Dias Ferreira, Larissa Xavier Neves da Silva, Angélica Trevisan De Nardi, Lucinéia Orsolin Pfeifer, Lucas Helal, Anderson Donelli da Silveira, Carisi Anne Polanczyk, Hirofumi Tanaka, Leonardo Alves, Leony Galliano, Linda S. Pescatello, Patrícia Martins Bock, Ruy Silveira Moraes, Beatriz D. Schaan, Stephanie Santana Pinto, Daniel Umpierre, The HAEL Study Group

## Abstract

**Importance:** Hypertension is a prevalent condition among the elderly, necessitating effective lifestyle interventions to reduce ambulatory blood pressure (BP) and improve overall health.

**Objective:** To determine whether a pragmatic combined exercise training program is more effective than a health education program in reducing ambulatory BP in older adults with hypertension.

**Design:** The “Hypertension Approaches in the Elderly: a Lifestyle study” (HAEL) was a multicenter, single-blinded, randomized clinical trial.

**Setting:** Participants were recruited from September 2017 and June 2020 in two centers in Brazil.

**Participants:** Individuals aged 60 years or older with a diagnosis of hypertension as assessed by ambulatory BP monitoring or current use of anti-hypertensive drugs were included.

**Interventions:** Participants (n=160) were randomized to a supervised, moderate-intensity, training (3 times/week) that involved aerobic and resistance exercises in a pragmatic setting or to a health education program including weekly, 1-hour workshops addressing hypertension management.

**Main outcomes and measures:** The primary outcome was the 3-month change in systolic BP assessed by 24-hour ambulatory BP monitoring. Secondary outcomes were diastolic BP, endothelial function, functional capacity, and quality of life. Data analyses were conducted using the intention-to-treat (ITT) and a prespecified per-protocol approach.

**Results:** Out of 717 individuals screened, 160 participants (mean [SD] age, 67.4 [5.5]; 104 females [62%]) were randomized. Compared with health education, exercise training did not reduce 24-hour ambulatory systolic BP (129.1 vs 128.9 mmHg; P=0.98) or diastolic BP (74.2 vs 74.4 mmHg; P=0.90), with measures remaining stable from baseline in both groups and no differences across daytime and nighttime timeframes. Compared with the health education program, combined exercise training increased peak oxygen consumption (0.4 ml/kg/min; 95% CI, -0.5 to +1.3) and Short Physical Performance Battery scores (0.7; 95% CI, +0.5 to +1.0). No major differences were detected between interventions for walking distance, quality of life, and endothelial function. The per-protocol analyses, restricted to non-dropout participants with ≥70% adherence to interventions, yielded results consistent with those obtained using the ITT approach.

**Conclusions and Relevance:** Among participants in the HAEL study over 3 months, combined exercise training was not superior to a health education program to reduce ambulatory BP.

**Trial Registration:** ClinicalTrials.gov Identifier: NCT03264443.

**Key Points:** **Question:** What is the comparative efficacy of a 12-week combined aerobic and resistance training versus health education in reducing ambulatory blood pressure among older adults with hypertension?

**Findings:** In this two-center randomized clinical trial including 160 older adults with hypertension, there were no significant changes in systolic or diastolic ambulatory blood pressure after 3 months of combined exercise training or health education program.

**Meaning:** A 12-week combined exercise training did not outperform health education in reducing ambulatory blood pressure in older adults with hypertension.

## Introduction

Hypertension affects over 60% of adults aged 55 years or older^1^, yet only 36% of older adults with hypertension meet physical activity guidelines^2^, despite exercise being an established treatment component^3^. This gap underscores the need for pragmatic, scalable interventions that integrate into healthcare systems while addressing the physiological changes and comorbidities of aging.

Combined aerobic and resistance exercise programs benefit overall health and can lead to blood pressure (BP) reductions in individuals with hypertension. However, there is insufficient evidence derived from studies specifically designed for older adults, especially through pragmatic interventions to facilitate implementation in public health settings ^4^. Additionally, there is a scarcity of trials employing the gold standard measure of BP monitoring, ambulatory or 24-hour BP. In a systematic review with meta-analysis of 68 trials that tested combined exercise interventions, only one study employed ambulatory BP monitoring^5^. Moreover, a systematic review of 391 randomized controlled trials indicates that the sample sizes are often limited to adequately produce reliable estimates of treatment.

We evaluated the effects of a 12-week combined exercise intervention on ambulatory BP in older adults with hypertension, compared with an active health education control group. We hypothesized that exercise would produce greater reductions in systolic ambulatory BP, and assessed secondary outcomes related to cardiovascular health and physical function.

## Methods

This randomized, single-blinded, multicenter, two-arm, parallel superiority trial was conducted in two cities in southern Brazil: Porto Alegre (coordinating center) and Pelotas. The trial was registered at ClinicalTrials.gov (NCT03264443), and the full protocol was previously published^6^. We followed the reporting recommendations from the Consolidated Standards of Reporting Trials^7^ and the Sex and Gender Equity in Research Guidelines^8^. *Participants, recruitment, and blinding*

Eligible participants were: 1) individuals aged ≥60 years with hypertension residing in or near the study cities; 2) stable pharmacological regimen for ≥4 weeks prior to enrollment; and 3) willingness to participate in either intervention. Exclusion criteria included recent cardiovascular events (<6 months), chronic heart failure (NYHA class III or IV), unstable arrhythmia, or other conditions precluding exercise program participation.

Randomization followed a 1:1 ratio with permuted blocks of random sizes, allocating participants to combined exercise training or health education. The allocation sequence was generated electronically and maintained by a researcher not involved in interventions or data collection. Outcome assessors were blinded to participant allocation.

From September 2017 to June 2020, recruitment employed a multi-faceted approach across both centers, including electronic health records, word-of-mouth referrals, print and electronic flyers, and media outreach. While both centers used these methods, each prioritized different strategies. Detailed recruitment results stratified by center, age, and sex are published elsewhere^9^.

### Interventions

Participants were allocated to follow a 12-week intervention consisting of a combined exercise training program or a health education program focusing on controlling hypertension. All participants answered a weekly survey on pharmacological treatment changes, adverse events, and exercise performed outside of the study.

### Combined exercise training

The participants allocated to combined exercise training intervention completed a supervised exercise program with individual sessions lasting approximately 1 hour three times a week. All sessions consisted of cardiorespiratory and muscular strengthening exercises and were structured with an initial warm-up (<5 min), followed by 20–30 min of aerobic exercise at moderate intensity and 2–3 sets of 4–5 multi-joint resistance exercises, emphasizing major muscle groups (lasting 15-20 min), and 5–10 min of cool-down. Exercises employed during the intervention are documented and publicly accessible (https://osf.io/56wrv/files - Exercise protocol)^10^. Cardiorespiratory exercise intensity was based on a rate of perceived exertion of 14-15 on the Borg’s rate of perceived exertion scale^11^. The participants achieved this target intensity walking or running (if needed). Resistance exercise intensities were monitored based on the rating of 4–8 (out of 10) of the OMNI perceived exertion scale^12^. The resistance exercises included multi-joint movement, using both upper and lower body exercises emphasizing daily-life activities such as sitting, standing up, pushing, and pulling. The exercise interventions utilized body weight and elastic band resistance, requiring minimal infrastructure and financial investment. This simplistic approach was designed to facilitate implementation in public health facilities lacking dedicated exercise spaces. Session structure, prescribed movements, timing, and progression were identical for both centers. To minimize differences between the intervention groups, all the participants in the exercise group received summarized oral information related to all topics of the health education intervention at the end of each exercise session.

### Health Education Program

Participants in the health education intervention attended an on-site weekly lecture lasting approximately 1 hour each, totaling 12 sessions in an attempt to control for attention from the investigative team. The education program was conducted in groups and was focused on standardized content delivered interactively by trained health professionals, approaching different themes related to hypertension management. Details about the educational program contents and additional details related to both intervention groups are in the study protocol^6^.

### Primary Outcome

The primary outcome was systolic ambulatory BP measured on the non-dominant arm for 24 hours with an automatic portable device (90207, SpaceLabs, Redmond, WA, USA). The final assessment was conducted at least 24 hours after the last intervention session and within 10 days after the participant completed the training intervention.

Participants were instructed to maintain a diary of daily activities and to avoid physically demanding and/or stressful activities while wearing the monitor. Ambulatory BP monitoring reports were considered valid if at least 70% of expected readings were available^13^.

### Secondary Outcomes

Office BP was measured using an oscillometric automatic device (OMRON Healthcare, Bannockburn, IL, USA) before and after interventions, following a similar timeframe to the ambulatory BP assessment. Participants rested quietly, seated comfortably for 5 min, with three measurements taken 1–2 min apart on the arm. The average of the three measurements was reported.

Cardiorespiratory fitness (VO_2_peak) was assessed through maximal cardiopulmonary exercise tests on a treadmill. Oxygen consumption, carbon dioxide production, and expired air volume were recorded breath-by-breath during an incremental protocol supervised by exercise physiologists and physicians. Initial speed was set at 2.0-2.5 km/h at 0% incline, with subsequent increases of 0.4-0.5 km/h and 1% incline every minute based on clinical history. Tests continued until subjective exhaustion or standard clinical stopping criteria. Different metabolic carts were used between centers (Porto Alegre: Quark CPET, COSMED, Italy; Pelotas: VO2000 MedGraphics, USA), though pre- and post-measurements used the same system at each center.

Flow-mediated dilatation was assessed using high-resolution ultrasonography (HD7XE, Phillips, USA). A high-frequency transducer (3-12 MHz) recorded brachial artery dilatation for 120 seconds following 5-minute cuff occlusion in fasted subjects. Flow-mediated dilation was calculated as the percentage change in arterial diameter from baseline to post-occlusion using Brachial Analyzer Software (Vascular Tools, USA).

A six-minute walk test was conducted as a secondary measurement of cardiorespiratory fitness. Following established guidelines^14^, participants were instructed to walk along a straight, enclosed path at a self-selected pace, aiming to cover as much distance as possible within six minutes. They received standardized verbal encouragement, and their final distance in meters was recorded for each test.

Health-related quality of life was assessed using the Brazilian Version of the Short Form 36 (SF-36), a 36-item survey covering eight domains: physical functioning, bodily pain, role limitations due to physical health problems, role limitations due to emotional problems, emotional well-being, social functioning, vitality (energy/fatigue), and general health. Scores for each domain are presented on a 0 (worst) to 100 (better) scale^15^.

The Short Physical Performance Battery (SPPB) was used to assess participants’ lower extremity function. This tool evaluates three key areas: balance, assessed by holding three progressively challenging standing positions; gait speed, measured by the time taken to walk 3 meters; and lower body strength, measured by the time to complete five chair stands. The scores from each component are then combined to yield an overall physical performance^16^.

### Descriptive Variables

Height and body weight were assessed using a stadiometer and calibrated scale as previously described ^12^. Waist circumference was assessed in the midpoint between the iliac crest and the 10th rib. Handgrip strength was measured with a mechanical hand dynamometer (Jamar Sammons Preston Rolyan, Bolingbrook, IL, USA) in both hands in an upright position and elbows flexed at 90°. The higher value of three attempts was considered as the maximal handgrip strength.

### Patient and public involvement

Patients and the public were not involved in the design, conduct, or reporting of this trial.

### Statistical Analyses

The sample size was estimated based on the systolic ambulatory BP response to exercise training and behavioral interventions. A sample of 184 subjects conferred adequate power to detect differences of 2.5 to 3.0 mmHg in the mean values of the 24-hour ambulatory BP monitoring for the primary outcome. More details on sample size estimation are available in the published protocol^6^.

Generalized estimated equations with an independence model as the covariance matrix and adjusted for baseline values were calculated for each variable of interest, with time and allocation group as factors. Post-hoc analyses were conducted, whenever applicable, using the Tukey test adjusted for multiple comparisons. Datasets were divided in intention-to-treat (ITT) - where all randomized individuals with at least one measurement (irrespective of adherence and completion status) were considered - and per protocol *-* where only non-dropout and adherent (≥70%) subjects were considered. Data are presented as estimated marginal means with 95% confidence intervals or as otherwise stated. Statistical significance was defined as a two-sided P<.05 for the primary and secondary outcomes.

Statistical analyses were performed using R version 4.2.2, running through RStudio 2023.06.1+524 “Mountain Hydrangea” Release (Posit Software, PBC, Boston, MA, USA). The main packages used in the analyses were ‘gee’ version 4.13-25 and ‘emmeans’ version 1.8.3. The full script for the analyses is public available (https://osf.io/56wrv/files)^10^.

### Protocol Deviations

The protocol had four main deviations. First, the Pelotas center closed due to low recruitment, affecting approximately half of its planned target. The coordinating center assumed remaining recruitment responsibilities. Second, the trial was terminated early due to COVID-19 restrictions in June 2020. After consulting external advisors, we ended the trial with 168 participants randomized and 160 completed. Third, the quality-of-life questionnaire was changed to the SF-36 mid-study to enable economic analyses, causing data to be missing for approximately 60 participants. Fourth, the autonomic function, a secondary outcome assessed via beat-to-beat BP variability, was not measured due to equipment failure.

## Results

A total of 168 participants were enrolled in the study. Eight participants were unable to finish the protocol. Therefore, data on 160 participants were analyzed. A flow diagram detailing the trial recruitment process is presented in **Figure 1**.

**Figure 1.**
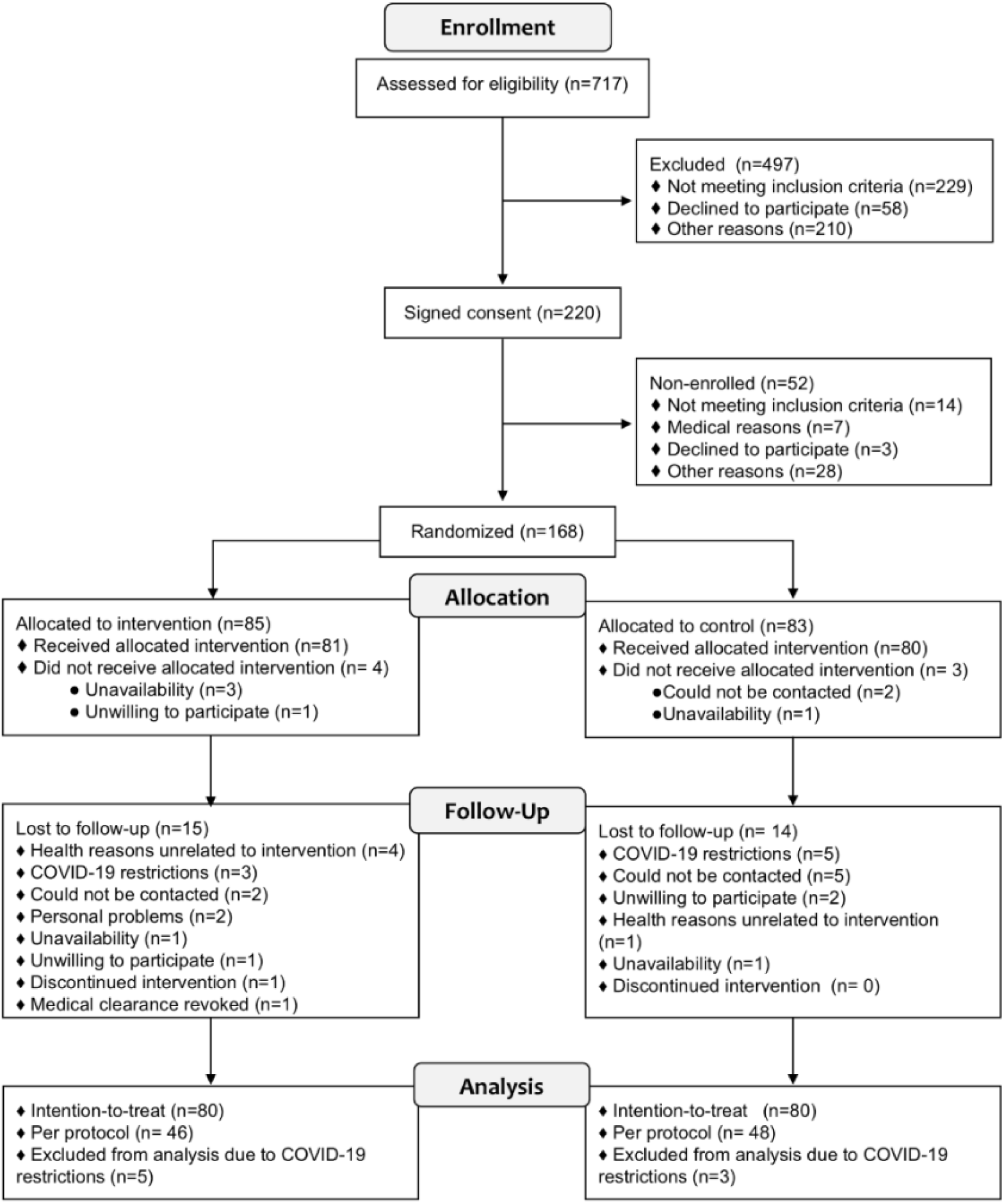
HAEL Trial consort diagram.

The mean age for both groups was approximately 67 ± 5.5 years. The majority of the sample consisted of women, individuals identifying as white, and participants with a body mass index marginally within the obesity range (**Table 1**). On average, BP was within the limits of hypertension control, with most participants receiving a combination of two or more anti-hypertensive medications. Adherence to the interventions was 52.5% for the combined exercise training group and 70.7% for the health education program in the intention-to-treat analysis. Among the participants who reached the per-protocol analysis threshold, adherence was 83.1% for the combined exercise training group and 88.4% for the health education program.

**Table 1.** Participant’s sociodemographic and clinical characteristics.

|  | **TOTAL** | **Combined exercise training** | **Health education program** |
| --- | --- | --- | --- |
| **Age, years (SD)** | 67.4 (5.5) | 67.5 (5.2) | 67.3 (5.9) |
| **Sex, n (%)** |  |  |  |
| Female | 104 (62%) | 56 (66%) | 48 (58%) |
| Male | 64 (38%) | 29 (34%) | 35 (42%) |
| **Race and Ethnicity, n (%)** |  |  |  |
| White | 131 (78%) | 62 (73%) | 69 (83%) |
| Black / Afro-descendants | 28 (17%) | 16 (19%) | 12 (15%) |
| Indigenous | 3 (2%) | 2 (2%) | 1 (1%) |
| Asian | 1 (1%) | 1 (1%) | 0 (0%) |
| Multiracial | 3 (2%) | 3 (4%) | 0 (0%) |
| Not reported | 2 (1%) | 1 (1%) | 1 (1%) |
| **Office BP, mean (SD), mmHg** |  |  |  |
| Systolic BP | 134 (20) | 134 (20) | 134 (20) |
| Diastolic BP | 79 (12) | 78 (13) | 79 (11) |
| **Antihypertensive medications, n (%)** |  |  |  |
| Beta-blockers | 70 (42%) | 39 (46%) | 31 (37%) |
| Angiotensin-converting enzyme inhibitors | 19 (11%) | 10 (12%) | 9 (11%) |
| Adrenergic antagonist | 20 (12%) | 10 (12%) | 10 (12%) |
| Calcium channel blockers | 46 (27%) | 21 (25%) | 25 (30%) |
| Diuretics | 36 (21%) | 17 (20%) | 19 (23%) |
| Combined | 114 (68%) | 60 (71%) | 54 (65%) |
| **Anthropometric, mean (SD)** |  |  |  |
| Waist circumference, cm | 103 (12) | 103 (12) | 102 (11) |
| BMI, kg/m<sup>2</sup> | 30.8 (5.0) | 30.8 (5.1) | 30.8 (4.9) |
| **Cardiopulmonary fitness, mean (SD)** |  |  |  |
| VO<sub>2</sub>peak (ml/kg/min) | 22.5 (5.5) | 21.8 (6.1) | 23.1 (4.8) |
| **Handgrip strength, mean (SD)** |  |  |  |
| Dominant arm, kg | 26 (11) | 26 (12) | 24 (10) |

Results express sociodemographic and clinical participants' characteristics at baseline for the total sample and stratified by intervention groups. SD: standard deviation; BP: blood pressure; BMI: body mass index; VO<sub>2</sub>peak: peak oxygen consumption.

### Primary and secondary outcomes

All estimated marginal means and p-values for interaction terms and pre-post post-hoc tests derived from the ITT dataset are presented in **Table 2**. Statistical analyses derived from the per-protocol dataset are presented in **Supplementary 1**.

**Table 2.** Changes in ambulatory BP monitoring and office BP.

| Timeframes and interventions | Baseline | 3 months | *p*-value interaction |
| --- | --- | --- | --- |
| Systolic BP in mmHg: Means (95% CI) |  |  |  |
| 24h ABPM |  |  |  |
| Exercise training (n=79) | 128.6 (128.2 - 129.1) | 129.1 (126.7 - 131.6) | 0.98 |
| Health education (n=79) | 128.4 (127.9 - 128.8) | 128.9 (127.0 - 130.8) |  |
| Daytime ABPM |  |  |  |
| Exercise training (n=79) | 130.9 (130.3 - 131.4) | 131.1 (128.4 - 133.7) | 0.57 |
| Health education (n=79) | 130.5 (129.9 - 131.1) | 131.8 (129.6 - 133.9) |  |
| Nighttime ABPM |  |  |  |
| Exercise training (n=79) | 122.9 (122.4 - 123.4) | 123.5 (120.8- 126.2) | 0.62 |
| Health education (n=79) | 122.7 (122.2 - 123.3) | 122.5 (120.0 - 125.0) |  |
| Office BP |  |  |  |
| Exercise training (n=80) | 135.1 (134.0 - 136.1) | 129.7 (126.2 - 133.1) | 0.45 |
| Health education (n=80) | 134.4 (133.3 - 135.6) | 131.0 (137.6 - 134.5) |  |
| Diastolic BP in mmHg: Means (95% CI) |  |  |  |
| 24h ABPM |  |  |  |
| Exercise training (n=79) | 74.6 (74.3 - 74.8) | 74.2 (72.8 - 75.7) | 0.90 |
| Health education (n=79) | 74.6 (74.4 - 74.9) | 74.4 (73.4 - 75.4) |  |
| Daytime ABPM |  |  |  |
| Exercise training (n=79) | 76.7 (76.4 - 77.0) | 77.0 (76.0 - 78.1) | 0.90 |
| Health education (n=79) | 76.6 (76.4 – 76.9) | 76.1 (74.5 - 77.7) |  |
| Nighttime ABPM |  |  |  |
| Exercise training (n=79) | 69.5 (69.1 - 69.8) | 69.2 (67.4 - 71.0) | 0.66 |
| Health education (n=79) | 69.5 (69.2 - 69.8) | 68.7 (67.1 - 70.3) |  |
| Office BP |  |  |  |
| Exercise training (n=80) | 78.7 (78.2 - 79.1) | 77.8 (75.9 - 79.7) | 0.81 |
| Health education (n=80) | 78.6 (78.2 - 79.1) | 77.4 (75.6 - 79.3) |  |

Results express baseline-adjusted estimated marginal means of blood pressure values, derived from intention-to-treat analysis. ABPM: Ambulatory blood pressure monitoring; BP: blood pressure.

### Blood Pressure

No significant between-group differences were observed for systolic or diastolic ambulatory BP across any timeframe (24-hour, daytime, or nighttime; all interaction P≥0.57). Within-group analyses revealed that 24-hour ambulatory BP remained stable in both groups from baseline to 3 months both in exercise training (systolic BP: 128.6 to 129.1 mmHg; diastolic BP: 74.6 to 74.2 mmHg) and health education participants (systolic BP: 128.4 to 128.9 mmHg; diastolic BP: 74.6 to 74.4 mmHg) (Table 2). Daytime and nighttime values similarly showed no changes.

For office BP, no significant between-group differences were detected (interaction P=0.45). However, the exercise training group showed a significant within-group reduction in systolic office BP from 135.1 mmHg (95% CI, 134.0-136.1) to 129.7 mmHg (95% CI, 126.2-133.1; 5.4 mmHg decrease, P<0.01), while diastolic office BP and both office BP measures in the health education group remained unchanged (Table 2). The per-protocol analysis yielded consistent findings.

### Functional Outcomes

For VO_2_peak, exercise training showed greater improvement than health education (exercise training: +0.4 ml/kg/min, 95% CI, −0.5 to 1.3; health education: −1.7 ml/kg/min, 95% CI −2.6 to −0.8; between-group difference: 2.1 ml/kg/min; interaction P=0.01). The six-minute walk test showed increases in both groups (P<0.01 and P=0.04 for exercise training and health education groups, respectively), with no statistically significant difference between them (p=0.08). Although between-group differences were not significant, within-group increases suggest potential walking gains from both interventions. The SPPB score increased significantly in the exercise training, with no significant change in the health education group.

In the per-protocol analysis, findings were consistent with ITT results for the six-minute walk test and SPPB outcomes but not for VO_2_peak, where no significant difference was observed. This supports the impact of the exercise intervention on functional capacity.

### Quality of Life and Vascular Function

No significant between-group differences were observed across SF-36 domains in ITT analyses. The health education group showed significant within-group improvements in “emotional” (P<0.01) and “vitality” domains (P<0.01), whereas the exercise group showed no changes in these domains. Both groups showed significant within-group improvements in “physical limitations” (P=0.04 for exercise training; P=0.03 for health education). No differences were detected in “general health”, “social functioning”, “physical functioning”, “pain”, or “mental health”.

Per-protocol analyses (Supplementary Material) revealed a significant between-group difference in “general health” (P=0.03) and a group*time interaction for “vitality” (P=0.04), favoring the health education group. Additional within-group improvements in the health education group were noted for “role-emotional limitation” (P<0.01) and “social functioning” (P=0.04). Other findings were consistent with ITT analyses.

No significant between-group differences were observed for flow-mediated dilatation at the mid-intervention and end-of-intervention assessments (Table 3).

**Table 3.**
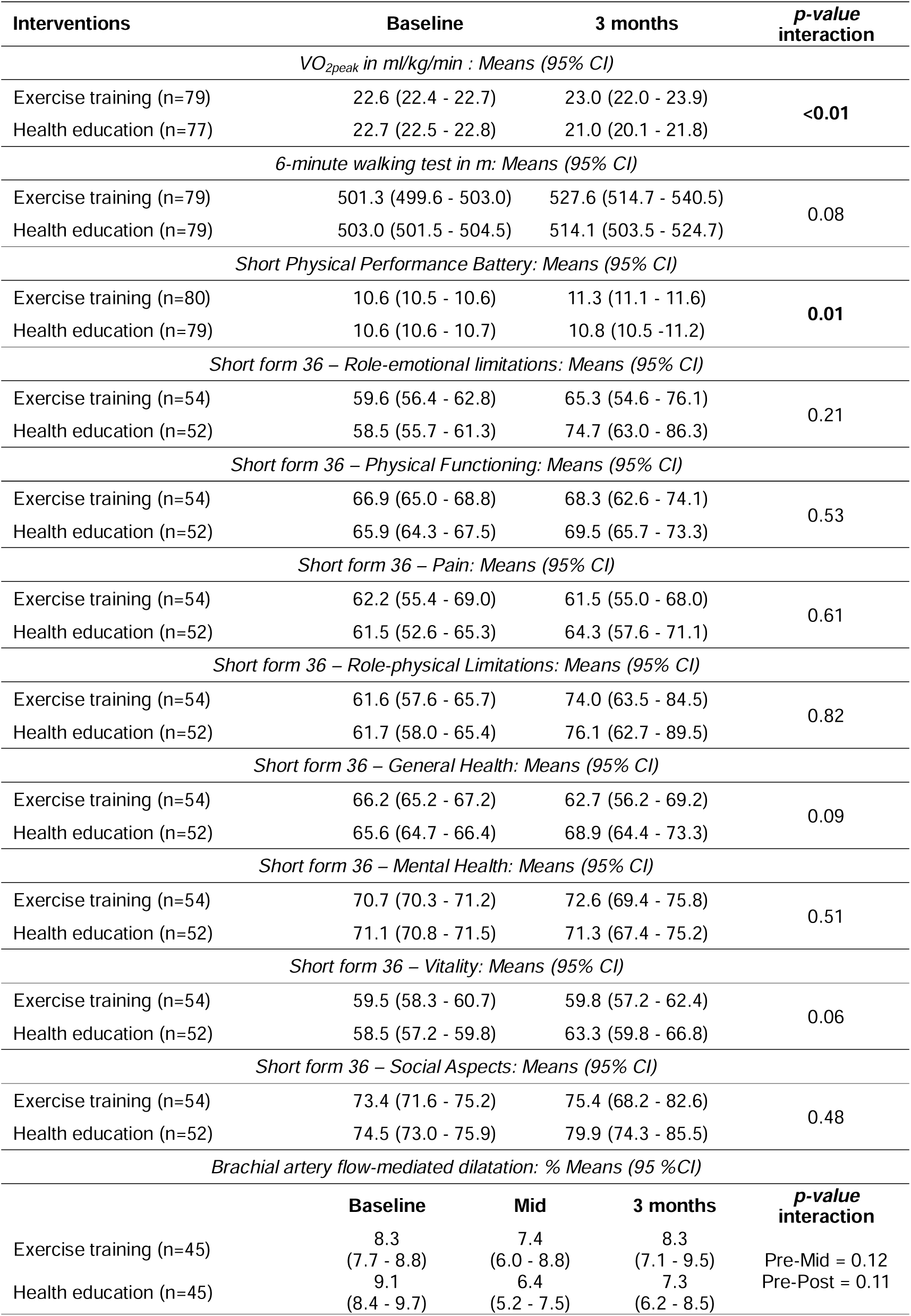
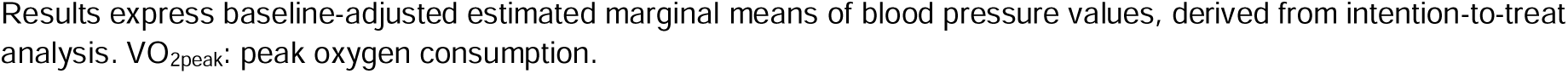
Changes in functional capacity, quality of life and endothelial function values. Changes in functional outcomes, SF-36 quality of life scores, and flow-mediated dilation.

| Interventions | Baseline | 3 months | p-value interaction |  |
| --- | --- | --- | --- | --- |
| VO<sub>2peak</sub> in ml/kg/min : Means (95% CI) |  |  |  |  |
| Exercise training (n=79) | 22.6 (22.4 - 22.7) | 23.0 (22.0 - 23.9) | <0.01 |  |
| Health education (n=77) | 22.7 (22.5 - 22.8) | 21.0 (20.1 - 21.8) |  |  |
| 6-minute walking test in m: Means (95% CI) |  |  |  |  |
| Exercise training (n=79) | 501.3 (499.6 - 503.0) | 527.6 (514.7 - 540.5) | 0.08 |  |
| Health education (n=79) | 503.0 (501.5 - 504.5) | 514.1 (503.5 - 524.7) |  |  |
| Short Physical Performance Battery: Means (95% CI) |  |  |  |  |
| Exercise training (n=80) | 10.6 (10.5 - 10.6) | 11.3 (11.1 - 11.6) | 0.01 |  |
| Health education (n=79) | 10.6 (10.6 - 10.7) | 10.8 (10.5 -11.2) |  |  |
| Short form 36 – Role-emotional limitations: Means (95% CI) |  |  |  |  |
| Exercise training (n=54) | 59.6 (56.4 - 62.8) | 65.3 (54.6 - 76.1) | 0.21 |  |
| Health education (n=52) | 58.5 (55.7 - 61.3) | 74.7 (63.0 - 86.3) |  |  |
| Short form 36 – Physical Functioning: Means (95% CI) |  |  |  |  |
| Exercise training (n=54) | 66.9 (65.0 - 68.8) | 68.3 (62.6 - 74.1) | 0.53 |  |
| Health education (n=52) | 65.9 (64.3 - 67.5) | 69.5 (65.7 - 73.3) |  |  |
| Short form 36 – Pain: Means (95% CI) |  |  |  |  |
| Exercise training (n=54) | 62.2 (55.4 - 69.0) | 61.5 (55.0 - 68.0) | 0.61 |  |
| Health education (n=52) | 61.5 (52.6 - 65.3) | 64.3 (57.6 - 71.1) |  |  |
| Short form 36 – Role-physical Limitations: Means (95% CI) |  |  |  |  |
| Exercise training (n=54) | 61.6 (57.6 - 65.7) | 74.0 (63.5 - 84.5) | 0.82 |  |
| Health education (n=52) | 61.7 (58.0 - 65.4) | 76.1 (62.7 - 89.5) |  |  |
| Short form 36 – General Health: Means (95% CI) |  |  |  |  |
| Exercise training (n=54) | 66.2 (65.2 - 67.2) | 62.7 (56.2 - 69.2) | 0.09 |  |
| Health education (n=52) | 65.6 (64.7 - 66.4) | 68.9 (64.4 - 73.3) |  |  |
| Short form 36 – Mental Health: Means (95% CI) |  |  |  |  |
| Exercise training (n=54) | 70.7 (70.3 - 71.2) | 72.6 (69.4 - 75.8) | 0.51 |  |
| Health education (n=52) | 71.1 (70.8 - 71.5) | 71.3 (67.4 - 75.2) |  |  |
| Short form 36 – Vitality: Means (95% CI) |  |  |  |  |
| Exercise training (n=54) | 59.5 (58.3 - 60.7) | 59.8 (57.2 - 62.4) | 0.06 |  |
| Health education (n=52) | 58.5 (57.2 - 59.8) | 63.3 (59.8 - 66.8) |  |  |
| Short form 36 – Social Aspects: Means (95% CI) |  |  |  |  |
| Exercise training (n=54) | 73.4 (71.6 - 75.2) | 75.4 (68.2 - 82.6) | 0.48 |  |
| Health education (n=52) | 74.5 (73.0 - 75.9) | 79.9 (74.3 - 85.5) |  |  |
| Brachial artery flow-mediated dilatation: % Means (95 %CI) |  |  |  |  |
|  | Baseline | Mid | 3 months | p-value interaction |
| Exercise training (n=45) | 8.3<br>(7.7 - 8.8) | 7.4<br>(6.0 - 8.8) | 8.3<br>(7.1 - 9.5) | Pre-Mid = 0.12<br>Pre-Post = 0.11 |
| Health education (n=45) | 9.1<br>(8.4 - 9.7) | 6.4<br>(5.2 - 7.5) | 7.3<br>(6.2 - 8.5) |  |

Results express baseline-adjusted estimated marginal means of blood pressure values, derived from intention-to-treat analysis. $\text{VO}_{2\text{peak}}$ : peak oxygen consumption.

### Adverse Events

In the exercise program group, a bone fracture was documented. This event occurred outside of the supervised exercise training sessions and, upon adjudication, was determined to be unrelated to the intervention protocol.

## Discussion

Contrary to our hypothesis, combined exercise training did not reduce ambulatory BP beyond the health education program. Although these findings differ from current evidence on exercise effectiveness for BP reduction, key methodological features distinguish the HAEL study from prior trials^17^.

The lack of ambulatory BP within- and between-group differences may stem from multiple factors. Our trial employed gold-standard 24-hour ambulatory BP monitoring, which provides a more accurate assessment of true BP levels due to measures less susceptible to white-coat effects at the beginning of interventions. In the office BP analysis, we observed a significant within-group reduction in the exercise training group, which presented a 5.4-mmHg reduction in systolic BP after the intervention (from 135.1 mmHg [95% CI, 134.0 – 136.1] to 129.7 mmHg [95% CI, 126.2 - 133.1]). Therefore, our results align with existing meta-analyses based on office BP measurements^18–20^.

Our sample presented adequate BP control at baseline. Given that responses to exercise interventions are related to initial BP values^21^, we cannot rule out a ceiling effect on BP responsiveness. This hypothesis is sustained by a meta-analysis evaluating BP responses to exercise interventions in a similar population as the one presented here, which showed that baseline BP was the strongest predictor of SBP and DBP reductions, explaining 74% and 53% of their variance, respectively^22^. This potential ceiling effect is also corroborated by the lack of ambulatory BP change in the health education program, which was developed based on an evidence-based strategy for BP reduction.

A key methodological strength in the HAEL study was the inclusion of an active attention control group, a feature often absent in most exercise-hypertension trials. Control participants underwent an evidence-based health education program focused on hypertension management. While this design choice enhances clinical relevance and controls for attention effects, neither the exercise nor education intervention altered ambulatory BP, suggesting factors beyond attention or behavioral engagement influenced our findings.

Exercise training improved VO2peak and SPPB scores but not six-minute walk distance, highlighting potential cardiovascular and disability risk reductions regardless of BP effects. In quality-of-life outcomes, both interventions improved “role-physical limitations”, suggesting that structured activities enhance self-perceived physical capacity. Health education showed advantages in “general health” and “vitality” (per-protocol analysis), possibly reflecting the less demanding intervention fostering favorable health outlook versus challenging exercise. These findings demonstrate complementary roles of exercise and education in improving older adult functional capacity and health perceptions.

## Strengths and limitations

Some limitations should be considered in our study. First, we did not systematically monitor changes in antihypertensive medication during the trial. Although we expect this was infrequent, this lack of specific control limit exploratory analysis of treatment responders. Second, the early termination of the study due to the COVID-19 pandemic may have affected the statistical power of our analyses. Although our power calculation with 160 participants was sufficient to detect differences as small as 2.5 mmHg for the primary outcome, we cannot rule out insufficient power for secondary outcomes. Third, challenges in scheduling end-of-study visits for some participants, though distributed evenly across groups, may have influenced BP effect sizes.

On the other hand, several characteristics of this trial enhance its real-world applicability and methodological rigor. A key strength is that the study was conducted in a low-resource, pragmatic setting that closely mimics real-life conditions common in low- and middle-income countries. Furthermore, the low-cost and scalable physical activity program was designed to capture the critical initial phase of older adults engaging with a new intervention. Such features, taken together with the use of an active control group and ambulatory BP monitoring, enhance both the rigor and generalizability of our findings to diverse healthcare settings.

## Conclusions

Combined exercise training was not superior to health education for reducing ambulatory or office BP in older adults with hypertension, despite improving cardiorespiratory fitness and lower limb function. Both interventions improved quality of life, highlighting the value of non-pharmacological programs. These findings underscore the complex relationships between exercise, BP control, and antihypertensive treatment in this population.

## Data Availability

All materials, data, and code are available online at Open Science Framework. Data for primary and secondary outcomes are embargoed until completion of peer review.

https://doi.org/10.17605/OSF.IO/56WRV

## Acknowledgements

The authors would like to acknowledge the role of Angelica Zanotto, SME-PMPA, and GPPG-HCPA staff members in supporting this research. Special thanks are given to individuals who have volunteered for this project.

