## Supplement - Tables 1 and 2 for "Combined Exercise Training vs Health Education for Older Adults with Hypertension: The HAEL Randomized Clinical Trial"

**Supplement 1.**

| Table 1. Changes in ambulatory BP monitoring and office BP derived from the per-protocol dataset. | | | |
| --- | --- | --- | --- |
| Timeframes and interventions | Baseline | 3 months | *p-value* interaction |
|  | *Systolic BP in mmHg: Means (95% CI)* | |  |
| 24h ABPM |  |  |  |
| Exercise training (n=46) | 129.6 (128.8 - 130.4) | 128.9 (126.2 - 131.7) | 0.82 |
| Health education (n=48) | 129.0 (128.3 - 129.6) | 128.7 (126.3 - 131.1) |  |
| Daytime ABPM |  |  |  |
| Exercise training (n=46) | 131.6 (130.7 - 132.5) | 130.8 (127.8 - 133.8) | 0.57 |
| Health education (n=48) | 130.8 (130.0 - 131.7) | 131.3 (128.7 - 133.9) |  |
| Nighttime ABPM |  |  |  |
| Exercise training (n=46) | 124.1 (123.5 - 124.8) | 123.8 (120.6 - 126.9) | 0.81 |
| Health education (n=48) | 123.7 (123.0 - 124.3) | 122.7 (119.7 - 125.7) |  |
| Office BP |  |  |  |
| Exercise training (n=46) | 136.0 (134.8 - 137.3) | 129.8 (125.8 - 133.7) | 0.23 |
| Health education (n=49) | 135.0 (133.7 - 136.2) | 132.1 (128.6 - 135.6) |  |
|  | *Diastolic BP in mmHg: Means (95% CI)* | |  |
| 24h ABPM |  |  |  |
| Exercise training (n=46) | 75.6 (75.3 - 76.0) | 74.3 (72.7 - 75.6) | 0.56 |
| Health education (n=48) | 75.7 (75.4 - 76.1) | 75.0 (73.8 - 76.3) |  |
| Daytime ABPM |  |  |  |
| Exercise training (n=46) | 77.5 (77.1 - 77.9) | 76.0 (74.2 - 77.8) | 0.21 |
| Health education (n=48) | 77.6 (77.2 - 78.0) | 77.7 (76.4 - 79.0) |  |
| Nighttime ABPM |  |  |  |
| Exercise training (n=46) | 70.7 (70.3 - 71.1) | 69.8 (67.9 - 71.8) | 0.61 |
| Health education (n=48) | 70.8 (70.4 - 71.2) | 69.2 (67.2 - 71.1) |  |
| Office BP |  |  |  |
| Exercise training (n=46) | 78.7 (78.1 - 79.4) | 76.7 (74.7 - 78.7) | 0.24 |
| Health education (n=49) | 78.8 (78.3 - 79.3) | 78.4 (76.5 - 80.3) |  |

Results express baseline-adjusted estimated marginal means of blood pressure values, derived from per-protocol analysis. ABPM: Ambulatory blood pressure monitoring; BP: blood pressure.

| Table 2. Changes in functional outcomes, SF-36 quality of life scores, and flow-mediated dilation derived from the per-protocol dataset. | | | | | |
| --- | --- | --- | --- | --- | --- |
| Interventions | **Baseline** | | **3 months** | | ***p-value* interaction** |
| *VO_2peak_ in ml/kg/min : Means (95% CI)* | | | | | |
| Exercise training (n=46) | 23.3 (23.0 - 23.5) | | 23.7 (22.5 - 24.8) | | **<0.01** |
| Health education (n=48) | 23.4 (23.1 - 23.6) | | 21.2 (20.3 - 22.1) | |  |
| *6-minute walking test in m: Means (95% CI)* | | | | | |
| Exercise training (n=46) | 500.0 (497.7 - 502.3) | | 527.4 (511.3 - 543.5) | | 0.11 |
| Health education (n=49) | 502.3 (500.5 - 504.1) | | 513.7 (502.2 - 525.2) | |  |
| *Short Physical Performance Battery: Means (95% CI)* | | | | | |
| Exercise training (n=46) | 10.7 (10.6 - 10.8) | | 11.4 (11.1 - 11.6) | | **0.01** |
| Health education (n=49) | 10.8 (10.7 - 10.8) | | 10.8 (10.5 -11.2) | |  |
| *Short form 36 – Role-emotional limitations: Means (95% CI)* | | | | | |
| Exercise training (n=31) | 60.9 (55.5 - 66.3) | | 66.6 (52.7 - 80.6) | | 0.15 |
| Health education (n=33) | 57.0 (51.8 - 62.2) | | 78.4 (64.6 - 92.1) | |  |
| *Short form 36 – Physical functioning: Means (95% CI)* | | | | | |
| Exercise training (n=31) | 71.4 (66.4 - 76.4) | | 72.9 (66.1 - 79.7) | | 0.61 |
| Health education (n=33) | 69.1 (64.6 - 73.6) | | 73.0 (67.3 - 78.6) | |  |
| *Short form 36 – Pain: Means (95% CI)* | | | | | |
| Exercise training (n=31) | 61.5 (53.2 - 69.8) | | 61.2 (53.3 - 69.2) | | 0.62 |
| Health education (n=33) | 58.8 (51.3 - 66.3) | | 62.5 (55.2 - 69.9) | |  |
| *Short form 36 – Role-physical Limitations: Means (95% CI)* | | | | | |
| Exercise training (n=31) | 59.1 (44.6 - 73.7) | | 72.0 (60.5 - 83.5) | | 0.85 |
| Health education (n=33) | 64.2 (50.8 - 77.6) | | 79.4 (66.7 - 92.2) | |  |
| *Short form 36 – General Health: Means (95% CI)* | | | | | |
| Exercise training (n=31) | 68.5 (66.5 - 70.5) | | 61.0 (52.0 - 69.9) | | **0.03** |
| Health education (n=33) | 66.1 (64.4 - 67.7) | | 70.4 (65.1 - 75.7) | |  |
| *Short form 36 – Mental Health: Means (95% CI)* | | | | | |
| Exercise training (n=31) | 72.3 (71.5 - 73.1) | | 75.8 (72.2 - 79.3) | | 0.50 |
| Health education (n=33) | 72.0 (71.2 - 72.8) | | 73.5 (68.8 - 78.1) | |  |
| *Short form 36 – Vitality: Means (95% CI)* | | | | | |
| Exercise training (n=31) | 61.6 (60.2 - 63.1) | | 60.8 (57.9 - 63.7) | | **0.04** |
| Health education (n=33) | 58.8 (57.2 - 60.5) | | 63.8 (59.6 - 68.0) | |  |
| *Short form 36 – Social functioning: Means (95% CI)* | | | | | |
| Exercise training (n=31) | 73.4 (71.6 - 75.2) | | 75.4 (68.2 - 82.6) | | 0.51 |
| Health education (n=33) | 74.5 (73.0 - 75.9) | | 79.9 (74.3 - 85.5) | |  |
| *Brachial artery flow-mediated dilatation: % Means (95 %CI)* | | | | | |
|  | **Baseline** | **Mid** | | **3 months** | ***p-value* interaction** |
| Exercise training (n=25) | 6.10 (5.38 - 6.83) | 5.03 (3.59 - 6.47) | | 6.72 (4.98 - 8.47) | Pre-Mid = 0.52  Pre-Post = 0.48 |
| Health education (n=31) | 6.65 (5.89 - 7.42) | 4.71 (3.31 - 6.11) | | 5.53 (4.43 - 6.64) |  |

Results express baseline-adjusted estimated marginal means of blood pressure values, derived from per-protocol analysis. VO_2peak_: peak oxygen consumption.
